# Rapid Scoping Review of Evidence of Outdoor Transmission of COVID-19

**DOI:** 10.1101/2020.09.04.20188417

**Authors:** Mike Weed, Abby Foad

**Author notes:** v.1.1 (10.7.2020) – not peer reviewed* | Protocol previously published on *medRxiv* https://doi.org/10.1101/2020.08.07.20170373.

## Abstract

The COVID-19 pandemic is both a global health crisis, and a civic emergency for national governments, including the UK. As countries across the world loosen their lockdown restrictions, the assumption is generally made that the risk of COVID-19 transmission is lower outdoors, and this assumption has shaped decisions about what activities can recommence, the circumstances in which they should re-commence, and the conditions under which they should re-commence. This is important for events and activities that generate outdoor gatherings of people, including both participatory and spectator sport events, protests, concerts, carnivals, festivals, and other celebrations.

The review, which was designed to be undertaken rapidly in 15 days, returned 14 sources of evidence of outdoor transmission of COVID-19, and a further 21 sources that were used to set the context and understand the caveats that should be considered in interpreting the review findings.

The review found very few examples of outdoor transmission of COVID-19 in everyday life among c. 25,000 cases considered, suggesting a very low risk. However risk of outdoor transmission increases when the natural social distancing of everyday life is breached, and gathering density, circulation and size increases, particularly for an extended duration. There was also evidence that weather had a behavioural effect on transmission, with temperatures that encourage outdoor activity associated with lower COVID-19 transmission. Due to lack of surveillance and tracing systems, and confounding factors and variables, there was no evidence that robustly tested transmission at outdoor mass gatherings (circa 10,000+ people), which are as likely to generate transmission from the activities they prompt (e.g. communal travel and congregation in bars) as from outdoor transmission at the gathering itself.

The goal of hosts and organisers of events and activities that generate outdoor gatherings of people is to prevent the escalation of risk from sporadic transmission to the risk of transmission through a cluster outbreak. Considerations for such hosts and organisers include: (1) does the gathering prompt other behaviours that might increase transmission risk?; (2) for each part of the event or activity, how dense is the gathering, how much do people circulate, how large is the gathering, and how long are people there?; (3) is rapid contact tracing possible in the event of an outbreak? These considerations should take place relevant to the size of the underlying risk, which includes the rate of infection in the community and the likely attendance of vulnerable groups. Risk must be balanced and mitigated across the risk factors of density, circulation, size and duration. No one risk factor presents an inherently larger risk than any other, but neither is any one risk factor a magic bullet to eliminate risk.

Finally, it is clear that the largest risks from gatherings come from spontaneous or informal unregulated and unmitigated events or activities which do not consider any of the issues, risks and risk factors outlined in this paper

## Introduction

Following 213 global deaths and 9,800 infections, on 30^th^ January 2020 the World Health Organisation categorised COVID-19 as a Public Health Emergency of International Concern (PHEIC), and five weeks later, on 11^th^ March, as a pandemic, at which point 118,000 cases and 4,291 deaths in 114 countries had been reported (Ghebreyesus, 2020). The first cluster of COVID-19 cases were recorded in Wuhan, China on 21^st^ December 2019, and the first death on 11^th^ January 2020. In the UK, the first domestically contracted case was recorded on 28^th^ February, and the first death on 5^th^ March. A global spread across Europe and North and South America meant that countries in every continent around the world, faced with a virus for which there was no vaccine and no treatment, implemented lockdown measures to deal with a global pandemic that had resulted in over 850,000 recorded deaths worldwide by 1^st^ September 2020 (Worldometer, 2020).

Following reductions in the spread of the virus as a result of these periods of lockdown, many countries are now developing and implementing plans to cautiously open up their societies and economies. Most of Europe has loosened lockdown measures and is now putting in place mitigation measures to live with the virus, including deciding which activities can re-commence, and which cannot. In the UK, after six weeks of lockdown, the government published a COVID-19 recovery strategy on 11^th^ May, with an update for further expansion published on 24^th^ July (HM Government, 2020a).

One of the assumptions in the UK recovery plan, and those of many other countries, is that the risk of transmission of the virus is lower outdoors (HM Government, 2020a). This assumption is central in informing decisions about which parts of the economy and society, and what activities, can re-commence, including decisions about the mitigation measures required. One area in which this is of particular importance is physical activity and sports, and particularly mass participation events such as running. Following government guidelines issued on 10^th^ July (HM Government, 2020b) it has been possible for mass participation running events to take place, and some have done so under principles drawing on government guidelines issued by UK Athletics (UK Athletics, 2020). These principles require additional mitigation measures to ensure some social distancing, including but not limited to, staggered wave starts, and strategies to limit pre-event gathering (such as not having on-site briefings, registration and announcements), and to accelerate post-event dispersal. However, for many event hosts, the issue of the risk of transmission of COVID-19 outdoors remains paramount, and this paper sets out a protocol for a rapid scoping review to explore evidence of such outdoor transmission. The findings of the review will also be relevant to events in other sectors that generate mass gatherings, such as concerts, carnivals and festivals.

## Review Scope

The review was designed to seek, evaluate and analyse evidence of incidents of outdoor transmission of COVID-19, the settings and environments of such transmission, and, where available, all relevant circumstances, including, but not limited to, temperature, wind conditions, social crowding or distancing, and the existence or otherwise of any COVID-19 mitigation measures. The review was also designed to seek, evaluate and analyse evidence of the prevalence of outdoor transmission compared to indoor transmission, and evidence of the impact of high profile mass gatherings, both immediately before (e.g. Champions League soccer matches) and during (e.g. Black Lives Matter protests) lockdowns.

While not designed to seek, evaluate or analyse evidence relating to the science of outdoor transmission of COVID-19, key insights from the extant science and literature, mostly identified as part of the review protocol, have been included to set the context and understand the caveats that should be considered in interpreting the review findings

The review was designed to be undertaken rapidly in 15 days. Therefore, a key additional purpose of the review was to assess whether a further extended more detailed and comprehensive review would capture wider and more extensive evidence.

## Review Protocol

The protocol for the review comprised three search elements: electronic searches using Google Scholar; pursuit of chains of sources referred to in papers returned in the electronic searches to the original source or sources of evidence; hand searches of papers and evidence sources considered by the UK government’s Scientific Advisory Group for Emergencies (SAGE), and its feeder groups, and of the research of known authors researching COVID-19 transmission.

The electronic searches were undertaken in Google Scholar, using the search string < “COVID 19” “outdoor” transmission >. Experimental searches suggested that “COVID 19” with a space within quotation marks would be the most effective search term, and that adding alternatives for COVID 19 (e.g. SARS-CoV 2, Coronavirus) added little to the efficacy of the search. Similarly, adding alternatives for “outdoor”, such as “outside” or “open air”, detracted from rather than improved search efficacy.

Search results were initially reviewed using the article title and preview text containing search terms returned by Google Scholar to evaluate whether a returned paper referred to outdoor transmission. If it appeared that it did, the full text of the paper was searched for the word “outdoor” and the relevant passages of text were reviewed to establish whether any evidence, or references to other sources of evidence, of outdoor transmission were included. Chains of sources referred to in papers were pursued to the original source or sources of evidence of outdoor transmission, which were also included for evaluation and analysis. Sources that provided opinion, summation or only onward reference to evidence of outdoor transmission were not included. Given the rapidly emerging evidence relating to COVID-19, neither peer-review status nor the outlet in which the source was published, was used as inclusion/exclusion criteria.

It was expected that evidence of outdoor transmission would be limited, and while such evidence was referred to in many sources, sources containing actual evidence were limited. For this reason, the initial search, ordered by Google Scholar’s relevance function, was limited to the first 100 sources returned because it was assumed that the search would be saturated at that point, and no new sources of evidence would be added. The pre-agreed marker of saturation was that the last 20 returns (81-100) would not add further sources of evidence, and this marker was met. Had this marker not been met, then the search would have been extended to the next 20 sources until the point was reached that the last 20 sources did not add any additional sources of evidence.

The search using Google Scholar’s relevance function was undertaken on 10/8/20 and resulted in 55 of the 100 returned sources being evaluated for inclusion. Of these 55 sources, 9 were included for joint review by the author team, as well as a further 12 from the pursuit of the chain of sources referred to in the 55 papers.

The search was then repeated, ordered by Google Scholar’s date added function. This was to ensure that new sources of evidence were not overlooked. The same protocol as described above was followed in relation to establishing the relevance of the returns, reference mining, and establishing the saturation point for the search, which was reached at 100 returns.

The search using Google Scholar’s date added function was undertaken on 11/8/20, and resulted in 28 of the 100 returned sources being evaluated for inclusion. Of these 28 sources, 2 were included for joint review by the author team, but none were added from the pursuit of the chain of sources referred to in the 28 papers.

In addition to the Google Scholar search, the papers and evidence sources considered by the UK government’s Scientific Advisory Group for Emergencies (SAGE), and its feeder groups, including the Scientific Pandemic Influenza Group on Modelling (SPI-M) and the New and Emerging Respiratory Virus Threats Advisory Group (NERVTAG), were hand searched for evidence and discussion of outdoor transmission on 13/8/20. Of these papers and evidence sources, 8 were included for joint review by the author team.

Finally, specific manual searches of the research of known authors researching COVID-19 transmission were undertaken during 10-16/8/20, of which 6 were included for joint review by the author team.

Overall, across all search elements, 37 sources were included for joint review by the author team. MW initially reviewed and evaluated sources for inclusion on the basis of relevance and was responsible for the decisions to include the sources for joint review by the author team as set out above. AF independently reviewed the inclusion decisions on the basis of relevance to the review. The author team then discussed the inclusions, and this resulted in the removal of two sources on the basis of relevance.

The author team also concluded that some of the included sources (14) provided direct evidence of incidents of outdoor transmission. However, others (21) did not provide evidence of incidents of outdoor transmission, but did provide insights, both from the science of transmission and the extant literature, that would be important in understanding the context and caveats that should be considered in interpreting the review findings. Consequently, the author team decided to include only the 14 sources providing direct evidence of outdoor transmission in the formal review, but to also develop a separate context and caveats discussion that would consider, but not be limited to, the remaining 21 sources. Figure 1 provides a summary decision flow of the search protocol and the inclusion decisions.

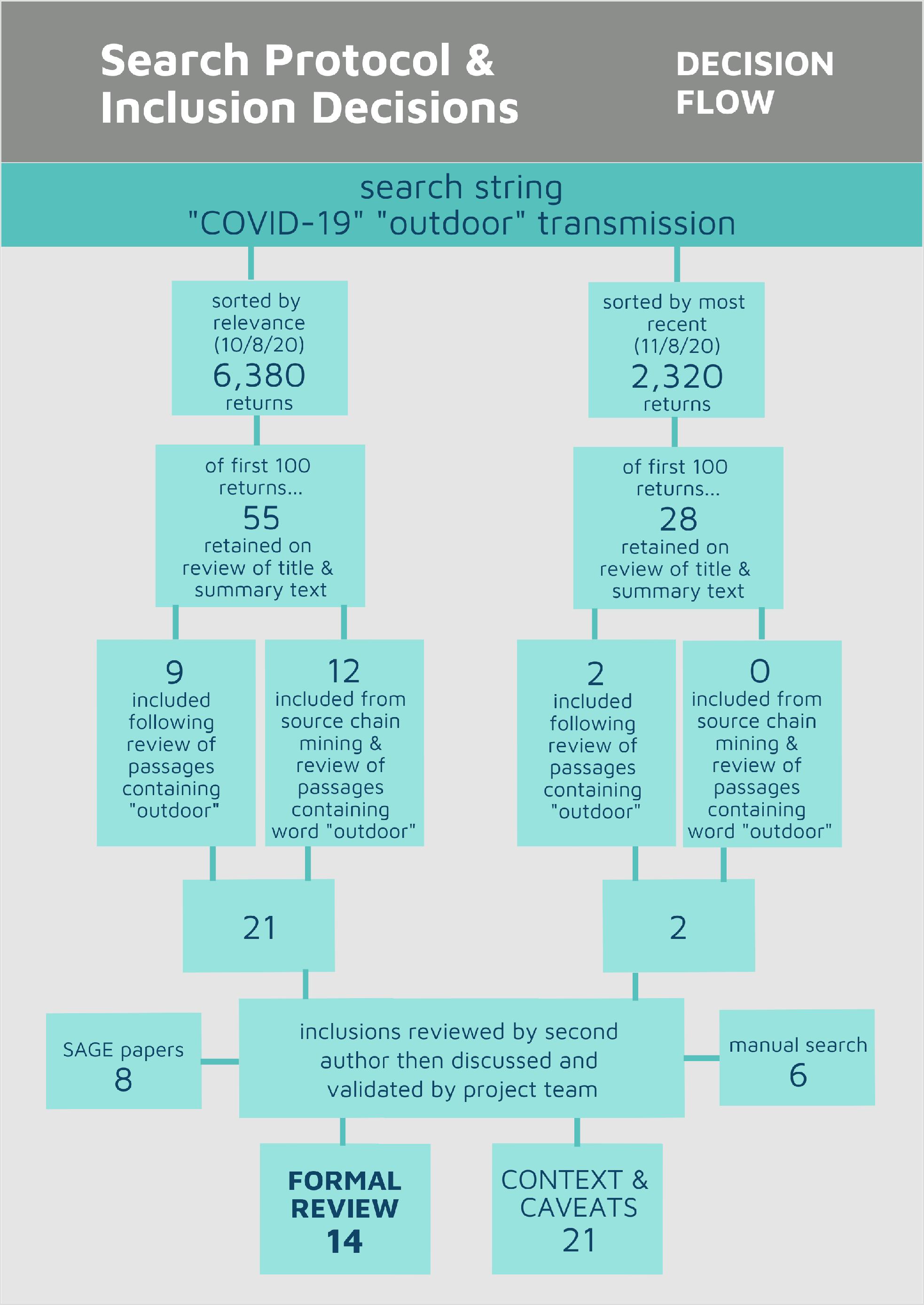
Figure 1

As evidence of incidents of outdoor transmission of COVID-19 was expected to be limited, search inclusion criteria related only to relevance – there were no inclusion/exclusion criteria related to quality or proxies for quality. However, quality was evaluated in the analysis and synthesis of the included sources. The product of this analysis is a critical *narrative synthesis* (Pope & Mays, 2006) which, whilst describing and synthesising evidence in substantive terms, also highlights potential weaknesses in returned evidence throughout the narrative.

MW wrote the first draft of the narrative synthesis, including an embedded evaluation of evidence quality. AF reviewed the evaluation of evidence quality embedded in the first draft of the narrative synthesis. The author team then discussed and agreed the embedded evaluation of evidence quality.

## Evidence of Outdoor Transmission of COVID-19: Review Findings

### What is the evidence of incidents of outdoor transmission?

The majority of the sources considered for inclusion in the review stated that transmission of COVID-19 outdoors is a lower risk than indoors. However, the evidence base for these statements can be traced back to just three root sources: a study of 110 cases in Japan (Nishiura et al, 2020), a review of case reports of 7,324 cases in China (Qian et al, 2020), and a database compiled at the London School of Hygeine & Tropical Medicine, totalling 20,471 cases across 616 clusters in the most recent publication (Lakha, Rudge & Holt, 2020), from which there had also been two earlier publications (Knight, Leclerc & Kucharski, 2020; Leclerc et al, 2020). Of these, only Leclerc et al (2020) has been peer reviewed.

Both Nishiura et al (2020) and Qian et al (2020) considered cases that largely occurred before any lockdown restrictions were imposed. The 110 cases in Japan were before 28/1/20, and Japan’s ‘state of emergency’ was imposed on 9/4/20. In China, the 7,324 cases were between 4/1/20 and 11/2/20, but all were outside Hubei province, where local lockdowns were variously introduced from 2/2/20. As such, the vast majority of cases occurred in ‘normal’ life when social interactions were unrestricted.

Nishiura et al (2020) present a very short, two page, unreviewed paper that does not provide details of primary data, nor of how the data was searched and extracted. It concludes that the likelihood of transmission in a closed environment is 18.7 times higher than outdoors, but it is not possible to validate the basis of this conclusion from the paper. It appears from a chart provided with the paper that, of the 110 cases, 11 (circa 10%) resulted in transmission to at least one other person in an outdoor environment, but only two of these created multiple cases, and none transmitted to more than three people. Whilst widely referenced in other papers, the findings of this paper should be treated with caution.

Of the 7,324 cases reviewed by Qian et al (2020), only one transmission incident, comprising two cases (a single transmission) took place in an outdoor environment, and was the result of a conversation between two individuals. The case reports were extracted from the health commissions of 320 municipalities in China (excluding those in Hubei province). Although this paper is unreviewed, and the data comprises only case reports, reasonable confidence can be placed in its findings, particularly as China has a comprehensive track and trace system.

The publications from the London School of Hygeine and Tropical Medicine (LSHTM) database comprise a peer-reviewed publication (Leclerc et al, 2020) and a report to SAGE (the UK government’s emergency scientific advisory group) (Knight, Leclerc & Kucharski, 2020) in June, and an unreviewed update published in July (Lakha, Rudge & Holt, 2020). The database is the result of a systematic search of peer-reviewed scientific reports of cases and media reports of cases. Following an earlier publication of the Leclerc et al (2020) paper on 1^st^ May, the authors sought to crowd source reports of further examples of transmission or outbreaks through a ‘suggested updates’ link on their publicly available database. The resulting updated publication (Lakha, Rudge & Holt, 2020) considered an estimated 20,471 reported cases across 616 clusters, and refined the categorisation of settings, which resulted in fewer settings being considered outdoor, or having an outdoor element. For example, in Leclerc et al (2020) and Knight, Leclerc and Kucharski (2020) religious settings were considered to have outdoor elements, but in Lakha, Rudge and Holt (2020) this had been refined to be indoor only. While the LSHTM database cannot be considered comprehensive, at over 20,000 cases it is extensive, and there is no reason to assume any bias in the reporting of indoor versus outdoor cases. Consequently, the findings can be treated with some confidence.

Of the 20,471 cases in the LSHTM database, only 461 in 11 clusters were associated solely with outdoor environments. A further 628 cases in 34 clusters were associated with environments that had some outdoor elements (Lakha, Rudge & Holt, 2020). As such, only 6% of cases, and 7% of clusters were associated with outdoor environments or environments with an outdoor element. 179 cases in 19 clusters were associated with sport environments, and a further 179 cases in 6 clusters were associated with parties, but in each case a significant majority of these were indoor. 100 cases in 6 clusters were associated with music venues, all of which were indoors.

Lakha, Rudge and Holt (2020) note that, despite lower risk of outdoor transmission, outdoor settings linked to crowding (e.g. markets and rallies) are linked to relatively large clusters (min 25 cases, max 163 cases), and in these settings people tend to circulate within the crowd. Similarly, numerous clusters were associated with close range interaction, and loud conversations, shouting or singing. Leclerc et al (2020) also note that duration in these contexts is important. Together this suggests that caution and further mitigation is likely to be required in relation to gathering density, circulation and size, as well as the duration of gatherings. Knight, Leclerc and Kucharski (2020) suggest that it is the interaction of environment, activity and duration that is important.

Two other sources referred to transmission in outdoor environments. One reported an outbreak at a summer camp where, in addition to outoor activities, participants shared dormitories and participated in singing (Szablewski et al, 2020). Another was a rapid review for consideration at SAGE in April (UNCOVER, 2020), which could find no high quality studies directly addressing the question of outdoor versus indoor transmission. Neither of the sources add to the insights above.

Across sources, there is limited evidence of transmission of COVID-19 in outdoor environments during the natural course of everyday life. Despite this limited evidence being found by only three sources, it collectively relates to over 25,000 cases with, in the cases of Qian et al (2020), Knight, Leclerc and Kucharski (2020), Leclerc et al (2020) and Lakha, Rudge and Holt (2020), extensive searches having been undertaken. Consequently, it is reasonable to assume that absence of evidence of extensive outdoor transmission of COVID-19 in everyday life can be taken to be evidence of absence of a meaningful risk of outdoor transmission.

However, there is some evidence to suggest that there is a higher risk of outdoor transmission in environments where the natural social distancing that takes place when ‘milling around’ in everyday life is breached, and gathering density, circulation and size is increased, particularly where this involves an extended duration. This could include aspects of outdoor concerts, festivals and some types of physical activity and sporting events.

### Has weather impacted transmission through encouraging indoor or outdoor activity?

Five included sources refer to a link between weather conditions and transmission, all of which associate lower temperatures with higher transmission. All sources suggest at least a partial role for a behavioural effect, in which lower temperatures encourage people to spend more time indoors. Four sources present independent analyses (Alvarez-Ramireez & Meraz, 2020; Carleton & Meng, 2020; Corripio & Raso, 2020; Newell, 2020), whilst one (UNCOVER, 2020) comments on the analyses presented by the first two. None of these sources have been peer-reviewed.

Carleton and Meng (2020) correlate global temperature data with a sample of 166,686 confirmed COVID-19 cases between 22/1/20 and 15/3/20 across 134 countries. Their results show a clear correlation between increased temperatures and reduced transmission, and they suggest three mechanisms: first, a biological effect on the virus itself; second, a behavioural effect, where people spend more time outdoors in less dense interactions; third, an increase in co-morbidities. Similarly, Alvarez-Ramireez and Meraz (2020) correlated daily cases in Wuhan, China between 29/1/20 and 6/3/20 with temperature trends, lagged for 6 days to account for the incubation period of the virus. This showed an increase in cases at temperatures below 8C (c.1,650 per day) and a decrease above 10C (c.350 per day), with this being attributed to human behaviour and an ‘indoor crowding effect’. UNCOVER (2000) concludes that the transmission effects demonstrated in these two papers are attributable to cool temperatures driving people indoors. However, a further exacerbating factor may be that people tend to have closer contact when gathering indoors than outdoors (‘indoor crowding’).

Corripio and Raso (2020) correlate meteorological data on temperature and humidity (lagged by 8 days) with COVID case records from the European Centre for Disease Protection and Control (for Italy) and the COVID-19 data Repository at John Hopkins University (for the USA) between 1/1/20 and 7/4/20, representing the time up to lockdown. Their data for Italy show a decrease in cases at temperatures over 11C, aligning with that found for Wuhan (Alvarez-Ramireez & Meraz, 2020). However, the USA data was unclear, although a very weak association between temperature and transmission was found. This led the authors to conclude that their data was consistent with poor outdoor transmission, and that any effect was more likely behavioural than biological.

Perhaps the most interesting evidence related to weather, which may also explain the unclear data from the USA found by Corripio and Raso (2020), is data correlating temperatures lagged by one week in six US states (California, Minnesota, Michigan, Ohio, Illinois, New York) with a ratio of the growth of cases between 10/3/20 to 18/6/20 (during periods of which some COVID-19 restrictions and mitigations were in place) (Newell, 2020). Newell’s (2020) analysis shows that infection rates in all six states fell distinctly at circa 50F (c.10C) and grew again from circa 70F (c.21C). Newell (2020) also presented two year average data showing that energy consumption of heating and airconditioning systems is lowest between 50F and 70F (c. 10C–21C). Together, this suggests that people switch off their heating and go outside when temperatures rise above 50F (10C), and come back inside again to utilise their air conditioning when the heat rises above 70F (21C). Newell (2020) concludes that lower infection rates are correlated with periods when people spend more time outdoors, and it may be that the second effect at 70F (20C) explains the lack of clear correlations in Corripio and Raso’s (2020) data.

Similar correlations between temperature and COVID cases have been found in each of the studies described, in which data sources and calculations are clear, and thus good confidence can be placed in these correlations, despite the lack of peer-review. It also appears reasonable to conclude that the correlations suggest at least a partial behavioural effect, where time spent outdoors leads to a fall in cases, and that this may be exacerbated by an indoor crowding effect where people tend to gather more densely in indoor settings.

A final possible effect to note is that if temperatures below 10C encourage people to spend more time indoors, this may have supressed, although likely no more than slightly, the numbers of cases of outdoor transmission found in the papers from Japan (Nishiura et al, 2020) and China (Qian et al, 2020) in the previous section, both of which experienced meteorological winter (December to February) during the period studied.

### What is the evidence for outdoor transmission of COVID-19 at mass gatherings?

‘Mass gatherings’, such as Champions League football matches in Italy, Spain and the UK, the Cheltenham Festival (A major UK horse racing event), and a rock concert in Cardiff (UK), have all featured in the media as potential drivers of COVID-19 transmission clusters that pre-date lockdowns. In addition, as lockdown restrictions have eased, media concerns have also been expressed about gatherings on beaches on sunny days and crowds generated by Black Lives Matter protests. However, only four sources of evidence relating to outdoor transmission at mass gatherings were found (Dave et al, 2020; Lazer et al, 2020; Sassano et al, 2020; and the LSHTM database, comprising Knight, Leclerc & Kucharski, 2020, Lakha, Rudge & Holt, 2020, and Leclerc et al, 2020), of which two (Sassano et al, 2020; Leclerc et al, 2020) were peer-reviewed.

Sassano et al (2020) present evidence on COVID-19 cases and deaths in Bergamo (Italy) following the Champions League football match between Bergamo’s Atalanta BC (the home team) and Valencia CF from Spain on 19/2/20. Although Atalanta were the home team, the game was held 35 miles away in Milan’s San Siro stadium due to its greater capacity -circa 45,000 fans attended the game, and it is estimated that circa 40,000 travelled from Bergamo.

There had been no recorded COVID-19 cases in Bergamo before the match, but Sassano et al (2020) present data showing that there were 1,815 cases three weeks after the game, and 8,803 cases and 2,060 deaths six weeks after the game. During March 2020, daily deaths in Bergamo were 568% higher than the average for the four years previous, compared to 187% higher in the wider Lombardy region. However, while it is possible that the occasion of the game may have contributed to this significantly higher number of ‘excess deaths’, attendance at the stadium is unlikely to have done so. The San Siro is an all seater stadium which, by design, holds each fan in a single place throughout the game, thus preventing them from circulating to any great extent with other fans. Sassano et al (2020) conclude that the interactions of Bergamo fans with each other and those outside their home town on transport, and in bars, clubs and other venues, including gatherings in homes, was likely to have been more significant for COVID-19 transmission than any outdoor transmission as part of the stadium crowd during the game itself.

Dave et al (2020) and Lazer et al (2020) each address the impact of Black Lives Matter protests in the USA, finding that the protests were either associated with no effect, or a reduction in the growth of COVID-19 cases. Lazer et al (2020) analyse 37,325 responses from two waves (12-28/6/20 and 10-26/7/20) of a monthly COVID-19 omnibus survey across 50 states, and show that there was a clear and significant negative correlation between the percentage of a state’s population that reported protesting, and subsequent COVID-19 cases. That is, the growth of COVID-19 cases was slower in those states where more people reported protesting. Lazer et al (2020) speculated that other mitigating behaviours may have been responsible for this, and showed that in states where more people reported protesting, adherence to mask wearing guidelines was higher.

A different explanation is offered by Dave et al (2020), who present data for 315 cities of more than 100,000 population that shows no difference in COVID-19 growth rates between cities that did (286) and did not (29) experience Black Lives Matter protests between 26/5/20 and 7/7/20.

However, Dave et al (2020) also draw on anonymised GPS data from mobile phone records to show a net increase in stay at home behaviour in cities that experienced protests. It is assumed that non-protestors stayed at home to avoid the perceived risk posed by the protests, and the data shows that the net impact was to increase the volume of social isolation, and thus prevent increased growth of COVID-19 cases, across the cities’ populations as a whole. Of course, it is possible that among protestors cases increased, but there is no data available to confirm or confound this.

The implications of the above in relation to outdoor transmission of COVID-19 is that there is no evidence that has meaningfully tested outdoor transmission at mass gatherings, and consequently an evaluation of evidence quality is superfluous. In each of the cases confounding factors are as likely, if not more likely, than outdoor transmission to explain upward, downward or neutral trends in COVID-19 case data. In addition, Leclerc et al (2020) note that, while a concert in Cardiff (UK), a horse-racing festival in Cheltenham (UK) and Champions League football matches in Italy, Spain and the UK that were the subjects of media concern could potentially have been connected to COVID-19 clusters, the absence of surveillance systems and rigorous testing means that no data is available and thus such connections remain speculation. The same is true of what appeared to be a crowded gathering of locals and tourists on Boscombe Beach (Bournemouth, UK) on 25/6/20, during which Bournemouth Council declared a ‘major incident’, asked people to stay away, and requested that police forces outside the area be put on alert to send reinforcements. Again, despite media interest, no testing or surveillance data is available.

It is clear, therefore, that there is an absence of evidence regarding outdoor transmission of COVID-19 at mass gatherings. However, outdoor mass gatherings are heterogenous, particularly in relation to crowd density, circulation and size, which largely comprise the activity element in the key interaction of environment, activity and duration that has been suggested to determine risk (Knight, Leclerc & Kucharski, 2020).

It is therefore also clear that absence of evidence in relation to mass gatherings cannot be assumed to be evidence that there is an absence of outdoor transmission at mass gatherings – this is likely to vary depending on the features and characteristics of different mass gatherings.

## Context and Caveats: Extant Science and Literature

Multiple sources exploring the science of transmission conclude that the risk of transmission of COVID-19 is low outdoors (Contini & Constable, 2020; Dominski & Brandt, 2020; TWEG, 2020; Redacted Author, 2020). Others conclude that aerosol transmission is likely to play a negligible role outdoors (Al Huraimel et al, 2020; EMG/NERVTAG, 2020), and with the risk of surface transmission being clearly identifiable from the features of venues (as well as being easily mitigatable), it is transmission by respiratory droplets that is the major area of concern for outdoor transmission (EMG, 2020a, 2020b).

In assessing the risk of outdoor transmission on COVID-19 as low, the science of transmission generally assumes that this risk is in the context of social distancing at or around 2m (ECDPC, 2020; EMG, 2020a; TWEG, 2020; NERVTAG, 2020), facilitated either by specific mitigation measures (Chu et al, 2020), or by the normal conventions of personal space and the more open outdoor environment that generally keeps groups of people further apart than indoors (Alvarez-Ramireez and Meraz, 2020; Slater, Christiana & Gustat, 2020). If these conventions are breached, then the extent of the breach in terms of proximity, duration, and circulation is significant. In this respect, TWEG (2020) conclude that the risk of outdoor droplet transmission in close, face-to-face contact in crowded areas is likely to be similar to that in some indoor settings.

Various studies suggest atmospheric conditions, such as air pollution, temperature and humidity (Contini & Constable, 2020; Sanchez-Lorenzo et al, 2020; Zoran et al, 2020), have a potential biological impact on COVID-19 transmission. However, as these studies did not consider any explanation other than biological mechanisms, we do not believe they contradict our review conclusions that there is at least a partial behavioural explanation related to lower COVID-19 transmission in temperatures when outdoor activity increases.

Reviews relating to the impact of mass gatherings on infectious disease transmission (McCloskey et al, 2020; Nunan & Brassey, 2020; Rainey, Phelps & Shi, 2016), which largely draw on pre-COVID-19 evidence, generally conclude that elevated risk comes with longer duration, crowdedness (particularly in communal overnight accommodation), and indoor environments. In terms of restrictions, Nunan and Brassey (2020) suggest that restricting mass gatherings closer to the epidemic peak may be more effective than restrictions applied further out. However, McCloskey et al (2020) and Nunan and Brassey (2020) each emphasise that, for COVID-19 transmission in particular, mass gatherings are not homogenous, and risks must be assessed on a case-by case basis.

Finally, some studies have explored the specific impact of particular activities. Following a number of reported outbreaks among choirs, NERVTAG (2020) concluded that the production of dropets during singing may be akin to a cough, although they noted there is no evidence describing the distance travelled by droplets during singing. They did recommend, though, that particular caution should be exercised with the environment (outdoor preferred), distancing (2m preferred), size and density (smaller groups preferred), arrangement (side-to-side rather than face-to-face) and duration (shorter preferred) when singing. These cautions apply not just to choirs, but to football crowds, and carnival, concert and festival go-ers, as well as to any other setting in which singing may take place.

In addition, some authors have speculated that exertion, such as that during physical activity and sport, may be similar to singing (Arias, 2020), while others have suggested similar cautions (Dominski & Brandt, 2020). But, while this seems intuitive, there are as yet neither incidents nor modelling studies that provide evidence for this. Similarly, a simulation study (Broken et al, 2020) has suggested a slipstream effect, in which walkers, runners and cyclists positioned directly behind the body width of another may need to distance for 5m, 10m and 20m respectively, or move slightly to the side, to avoid droplets. However, when SAGE considered evidence of extended carry due to downwind flow conditions (Redacted Author, 2020) they concluded that this was unlikely to be a significant route for infection unless people are in position for a long period of time and presumably unable to move. This suggests that the general considerations for duration and density set out in other contexts are the key mitigations that should apply.

These insights from the science of transmission and the extant literature align with our review evidence that the risk of outdoor transmission is low, unless the natural social distancing that takes place when ‘milling around’ in everyday life is breached. In such circumstances, caution and further mitigation is likely to be required in relation to gathering density, circulation, size and duration of contact(s). Knight, Leclerc & Kucharski (2020) suggest that the interaction of environment, activity and duration is a useful framework to assess risk and relevant mitigations which, importantly, will not be homogenous across different outdoor activities, gatherings, events and environments. However, outdoor events and activities are heterogenous, particularly in relation to gathering density, circulation and size, which largely comprise the activity element in the key interaction of environment, activity and duration that has been suggested to determine risk.

## Considerations for Events and Activities that Generate Outdoor Gatherings

The World Health Organisation describes four transmission scenarios for COVID-19: no cases, sporadic cases, clusters of cases, and community transmission (WHO, 2020). When countries or areas enter lockdowns, it is because cases have progressed to *community transmission*, where large numbers of cases not linkable to transmission chains exist. However, as countries or areas exit lockdowns, it is assumed to be because community transmission has been brought under control, and that cases can now be traced to transmission chains or clusters. In this scenario, given that achieving *no cases* is unlikely without a vaccine, continued isolated *sporadic cases* of transmission are inevitable, and so the key goal is to prevent transmission through *clusters of cases* focused in a particular location and/or activity. It is this latter risk that should be considered by hosts and organisers of events and activities that generate outdoor gatherings.

A framework for assessing this risk that focuses on the environment, activity and duration of a gathering has been suggested by Knight, Leclerc and Kucharski (2020). Both this review and the extant science and literature agree that an outdoor environment presents a low risk due to the natural social distancing that happens through the normal conventions of personal space in everyday life, as well as the wider open space of the outdoors that generally keeps groups of people further apart than indoors. Consequently, the key risks to focus on are those related to the activities and their duration that take place at outdoor gatherings where this natural social distancing might be breached.

In both the review and the extant science and literature, density, circulation, size and duration of the gathering generated by the activity or event have been highlighted as key risk factors.

The *density of the gathering* relates to the closeness of contact between individuals, and includes their arrangement (e.g. face-to-face or side-by-side). NHS Track and Trace defines ‘close contact’^1^ as spending more than 15 minutes within two metres of someone who has tested positive for COVID-19, or having face-to-face contact within one metre, or having skin contact, or being within one metre of an infected person for more than one minute without face-to-face contact.

*Circulation within the gathering* refers to how far individuals move around and bring themselves into contact with a larger number of other people. For example, in an all-seater football stadium or at the start of a running race, people ususally maintain their position in relation to those around them: at a festival or carnival, people often move around within the gathering and come into contact with a larger number of people.

The *size of the gathering* relates to the likelihood that infected individuals or groups might be present. Neither the review nor the exploration of extant science and literature has considered factors such as the underlying infection rate in a community, or case fatality rates, which were beyond the scope of the review. However, the baseline underlying risk will be lower in countries and communities where the underlying rate of infection is low, and the risk of the size of gathering should be considered in relation to this.

The *duration of the gathering* relates both to the gathering as a whole (e.g. a multi-day festival; an afternoon football match) and to individual elements of it (e.g. standing in a crowd, queing at the bar, visiting the toilet; before, at the start, during, and after a running race).

No one risk factor presents an inherently larger risk than any other, and all of the risk factors mitigate each other. For example: a more dense gathering is mitigated if circulation or duration is low; a larger gathering is mitigated if it is less dense, or if less time is spent in it; a longer duration is mitigated at lower levels of gathering density, size or circulation; and so on. Importantly, risk factors must be considered in relation to the size of the underlying risk, comprising elements such as infection rates in the community, the extent to which vulnerable or susceptible groups might be present, and the indoor or outdoor nature of the gathering. If the *density* of an outdoor gathering is such that the natural social distancing that happens through the normal conventions of personal space in everyday life is not breached, then the underlying risk is largely mitigated. However, if *density* is increased beyond the norms of natural social distancing and personal space, then risk will need to be mitigated across the other risk factors of *circulation, size* and/or *duration*. Risk might be mitigated extensively by one or two risk factors, or moderately across three or four, either to offset any increases in other risk factors, or to mitigate a larger underlying risk. Ultimately, the task for hosts and organisers is to achieve a balance across risk factors that will mitigate the underlying risk, and avoid a *cluster outbreak*.

Complicating matters further, the risk factors should be considered in relation to each aspect of an event or activity that has different elements. Risk should be considered for each individual aspect, and in aggregate for the event or activity as a whole.

In addition to the above, there are two other factors that should be considered by hosts and organisers of events and activities that generate outdoor gatherings of people. Firstly, the review highighted the potential for events and activities to prompt other behaviours that might be higher risk than the gathering itself. These include communal travel to the event or activity, indoor congregation in bars, cafes or other venues, and collective stays in overnight accommodation. Secondly, a key mitigation to prevent *cluster outbreaks* generating secondary cases (i.e. subsequent transmission away from the event or activity) and heightening the risk of elevation to *community transmission*, will be the ability to rapidly trace attendees and contacts at the event or activity. These issues are integrated with the discussion of risk factors above, and summarised in Figure 2.

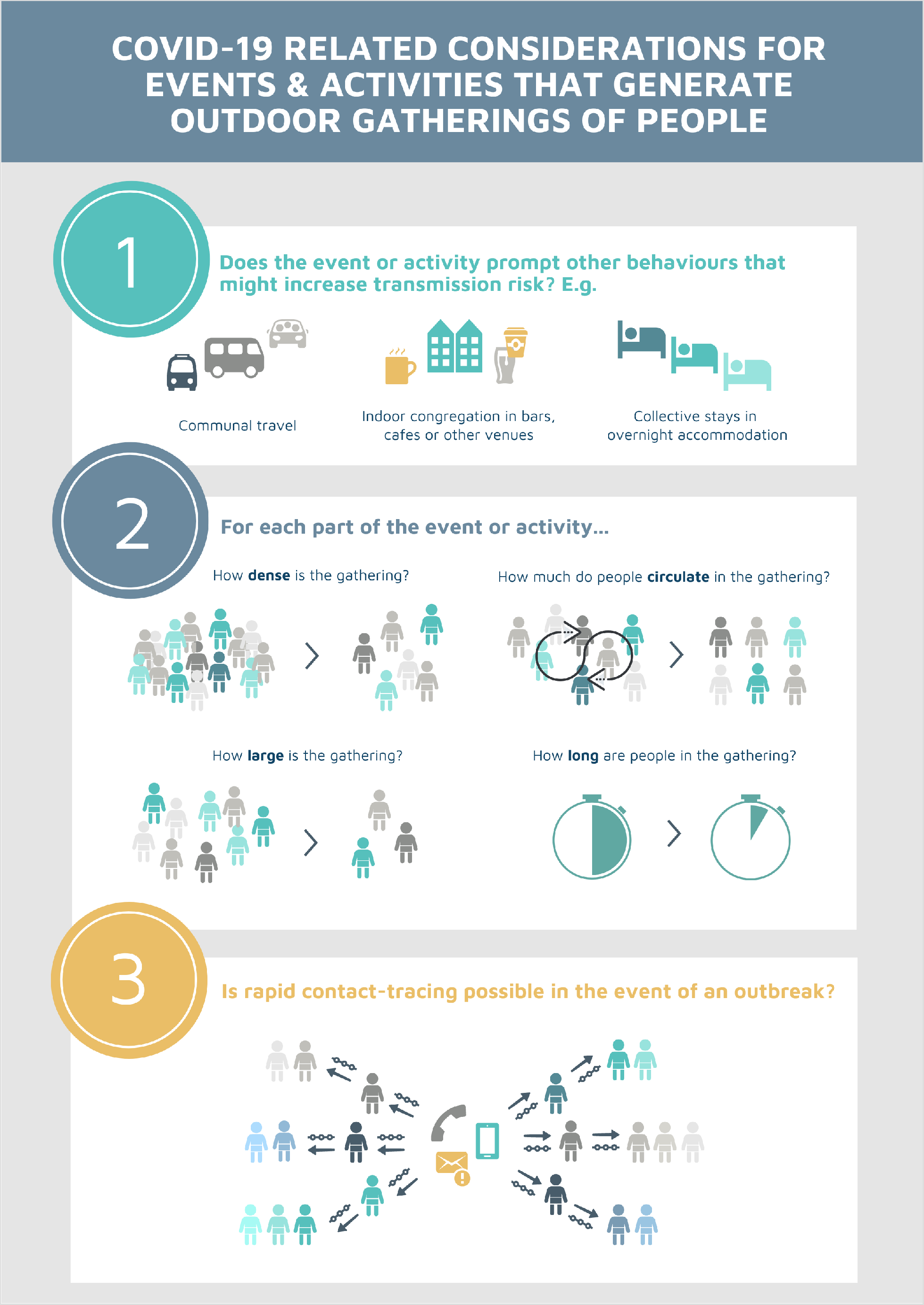
Figure 2

The considerations presented here relate to hosting or organising an event or activity. However, there are a three other wider factors that might be considered. Firstly, perhaps the largest risks from gatherings come from spontaneous or informal unregulated and unmitigated events or activities which do not consider any of the issues, risks and risk factors outlined in this paper (WHO, 2020). While attempts could be made to ban or regulate such events or activities, providing gatherings that have been properly risk assessed would be a mitigating factor against spontaneous or unregulated gatherings. Secondly, and related, given that many events and activities have cultural, social, economic and even political implications (WHO, 2020), the opportunity cost of not hosting or organising the activity or event should be considered. This could be a rise in riskier unregulated activity, or a fall in physical and mental health and wellbeing that could have wider morbidity and mortality impacts. Thirdly, the public appetite or aversion for events and activities that generate outdoor gatherings should be considered, as should the media coverage that drives this. This will also relate to what other events and activities are and are not taking place. Honey-Roses et al (2020) and Slater et al (2020) each address aspects of this issue, with Honey-Roses et al (2020) noting that there may be a longer term impact on our perceptions of outdoor spaces, their carrying capacity, and their design. For example, will public perceptions of what is crowded change, and will health criteria (both to prevent infection and promote healthy behaviours and activities – e.g. widening footpaths and trails) be mainstreamed into the design of outdoor spaces? And will such infrastructural changes be expected as part of the continued re-opening of society, or in the longer term?

## Conclusion and Limitations

The protocol for this review was designed to allow rapid completion in 15 days. This is clearly a limitation. However, the review was also designed to scope the available evidence. Both electronic searches were saturated within 100 returns, and the same root sources of evidence underpinned the majority of sources reviewed and evaluated for inclusion. Therefore, while it is possible that individual sources of relevant evidence may not have been captured by the review, we consider it highly unlikely that any further significant bodies of evidence relevant to this review were available when the searches concluded on 16/8/20. A more substantial review, with a more detailed and extensive search strategy, is therefore unlikely to be a productive enterprise.

Available evidence of outdoor transmission of COVID-19 has been reviewed, and the context and caveats provided by the extant science and literature considered. This leads to the conclusion that the outdoor environment presents a low risk of transmission of COVID-19 due to the natural social distancing that happens through the normal conventions of personal space in everyday life. However, the areas in which risk increases when the normal conventions of personal space are breached in outdoor environments have also been discussed and outlined, and these have been translated into considerations for hosts and organisers of events and activities that generate outdoor due to the natural social distancing that happens through the normal conventions of personal space in everyday life. However, the areas in which risk increases when the normal conventions of personal space are breached in outdoor environments have also been discussed and outlined, and these have been translated into considerations for hosts and organisers of events and activities that generate outdoor gatherings, and discussed how risks can be balanced. It is, of course, the balance of such risks that must be considered as, almost by definition, no gathering (even that within households) can be low risk in all areas.

## Data Availability

Data comprises the sources of evidence returned in the review search. All are publicly available and listed in the reference list.

## Conflict of Interests

MW is a member of the parkrun International Research Board, and regularly takes part in parkrun.

## Funding

This work is funded by parkrun.

## Footnotes

1 https://www.gov.uk/government/publications/guidance-for-contacts-of-people-with-possible-or-confirmed-coronavirus-covid-19-infection-who-do-not-live-with-the-person/guidance-for-contacts-of-people-with-possible-or-confirmed-coronavirus-covid-19-infection-who-do-not-live-with-the-person (accessed 30/8/20)

## Notes

### Competing Interest Statement

MW is a member of the parkrun International Research Board, and regularly takes part in parkrun. This work is funded by parkrun.

### Funding Statement

The work is funded by parkrun

### Author Declarations

Ethical approval, which gave an exemption from full review following a proportionate review, was provided by the Ethics Panel of the Faculty of Science, Engineering and Social Science at Canterbury Christ Church University. Ref: ETH2021-0004

### Summary of Updates

Error corrected in Conclusions and Limitation section in which several sentences of text were mistakenly repeated.

